# Burden of diarrhea and antibiotic use among children in low-resource settings preventable by *Shigella* vaccination: a simulation study

**DOI:** 10.1101/2023.07.03.23292159

**Authors:** Stephanie A Brennhofer, James A Platts-Mills, Joseph A Lewnard, Jie Liu, Eric R Houpt, Elizabeth T Rogawski McQuade

## Abstract

**Background:** *Shigella* is a leading cause of diarrhea and dysentery in children in low resource settings, which is frequently treated with antibiotics. The primary goal of a *Shigella* vaccine would be to reduce mortality and morbidity associated with *Shigella* diarrhea. However, ancillary benefits could include reducing antibiotic use and antibiotic exposures for bystander pathogens carried at the time of treatment, specifically for fluoroquinolones and macrolides (F/M), which are the recommended drug classes to treat dysentery.

**Methods:** We used data from the Etiology, Risk Factors, and Interactions of Enteric Infections and Malnutrition and the Consequences for Child Health and Development (MAL-ED) study to estimate the impact of two one-dose (6 or 9 months) and three two-dose (6 & 9 months, 9 & 12 months, and 12 & 15 months) *Shigella* vaccines on diarrheal episodes, overall antibiotic use, and F/M use. Further, we considered additional protection through indirect and boosting effects. To estimate the absolute and relative reductions in the incidence of diarrhea and antibiotic use under each vaccination scenario, Monte Carlo simulations with random sampling were performed.

**Findings:** We analyzed 9392 diarrhea episodes and 15697 antibiotic courses among 1715 children in the MAL-ED birth cohort study. There were 273.8 diarrhea episodes, 30.6 shigellosis episodes, and 457.6 antibiotic courses per 100-child years. A *Shigella* vaccine given at 9 & 12 months prevented 1.7 (95% CI: 1.3, 2.1) severe *Shigella* diarrhea episodes (46.5% reduction), 11.0 (95% CI: 10.0, 11.9) *Shigella* diarrhea episodes of any severity (35.9% reduction), 3.1 (95% CI: 2.6, 3.7) F/M courses (2.9% reduction overall), 5.8 (95% CI: 5.2, 6.6) antibiotic courses (1.0% reduction overall), and 6.3 (95% CI: 5.2, 7.5) F/M (3.2% reduction) and 11.2 (95% CI: 9.7, 12.9) antibiotic (1.2% reduction) exposures to bystander pathogens, respectively, per 100-child years.

**Interpretation:** A *Shigella* vaccine could make substantial reductions in *Shigella* diarrhea, antibiotic use to treat shigellosis, and bystander exposures due to shigellosis treatment. However, the reductions in overall diarrhea episodes and antibiotic use would be modest.

**Funding:** Wellcome Trust, Bill & Melinda Gates Foundation

## INTRODUCTION

*Shigella* is a leading cause of diarrhea and dysentery in children under the age of five in low- and middle-income countries (LMICs)^1^. In the multisite Etiology, Risk Factors, and Interactions of Enteric Infections and Malnutrition and the Consequences for Child Health and Development (MAL-ED) cohort study, *Shigella*-attributed diarrhea was found to have an incidence of 26.1 episodes per 100 child years in the first two years of life.^2^ There are several *Shigella* vaccines in the pipeline, of which three are in phase IIA and one in phase III trials.^3^ The World Health Organization (WHO) recently published preferred product characteristics (PPC) for a *Shigella* vaccine,^1^ and efforts are underway to define the full value proposition for such a vaccine. The primary goal of a *Shigella* vaccine is to prevent mortality and moderate-to-severe episodes of shigellosis with an efficacy target set by the WHO of 60% or more. Assuming this target can be met in trials conducted in ideal settings, real world estimates of the reduction in diarrhea episodes that would be expected after vaccine introduction are needed to predict population-level vaccine impact.

Furthermore, a *Shigella* vaccine may produce ancillary benefits that need to be quantified, specifically reductions in antibiotic exposures since diarrhea is a major cause of antibiotic use.^4, 5^ Previous analyses have identified *Shigella* as a leading contributor to antibiotic consumption among children in low-resource settings.^5^ In MAL-ED, *Shigella* was responsible for 14.8 antibiotic courses per 100-child years.^4^ Furthermore, 20.9% and 16.2% of all fluoroquinolone and macrolide courses given for diarrhea, respectively, were to treat shigellosis.^4^

Vaccine impact on fluoroquinolone/macrolide (F/M) use is of particular interest as they are the recommended treatment by the WHO for dysentery,^6, 7^ and 34% of dysentery cases in children under two years of age in the MAL-ED birth cohort were attributed to *Shigella*. Frequent use of antibiotics drives selection for drug resistant pathogens,^8^ and drug-resistant shigellosis is of concern.^6^ Azithromycin and fluoroquinolone resistant strains of *Shigella* are common in Asia and are growing in prevalence elsewhere.^9–11^

In addition to preventing exposures to antibiotics for *Shigella,* a *Shigella* vaccine could further reduce selective pressure on asymptomatic enteric pathogens (i.e., bystander pathogens) present in the gut at the time of shigellosis treatment. Bystander pathogens are not the target of treatment, but nonetheless are still exposed to antibiotics and are therefore at risk for development of antimicrobial resistance (AMR). There were more than 7 antibiotic exposures per child-year for bystander enteropathogenic bacteria in MAL-ED.^5^

To inform the vaccine value proposition, we aimed to quantify the potential impact of a *Shigella* vaccine on the incidence of *Shigella* diarrhea (severe and non-severe), all diarrhea, and antibiotic use in the first two years of life via various potential vaccination strategies. We considered different vaccine efficacies, dosing schedules, and types, including leaky vaccines (i.e., prevention of a fraction of episodes in all children) and all-or-nothing vaccines (i.e., prevention of all episodes in a subset of children who are vaccine responders).^12, 13^ We also quantified the potential impact of indirect protection to children who were too young to be vaccinated and the impact of a vaccine that performs better for children who have been previously exposed to *Shigella*.

## METHODS

### Study design and participants

The MAL-ED study design has been previously detailed.^14^ Briefly, this study was conducted at eight sites (Dhaka, Bangladesh; Fortaleza, Brazil; Vellore, India; Bhaktapur, Nepal; Loreto, Peru; Naushero Feroze, Pakistan; Venda, South Africa; and Haydom, Tanzania) from November 2009 to February 2014. Children were enrolled within 17 days of birth and followed for two years. Fieldworkers conducted twice weekly home visits to collect information on daily antibiotic use and presence of illness. Stool samples were collected monthly (non-diarrheal surveillance samples) and during diarrheal episodes. Diarrhea episodes were defined as three or more loose stools in a 24-hour period or the presence of blood in at least one stool. Diarrhea severity was determined by the modified Vesikari score, previously outlined.^15^

### Stool testing

The QIAamp Fast DNA Stool Mini Kit (Qiagen) was used to extract total nucleic acid from the stool specimens.^16^ To detect the presence of 29 enteropathogens via quantitative polymerase chain reaction (qPCR), TaqMan Array Cards (TAC) were run using AgPath One Step RT PCR kit (Thermo-Fisher).^2^ The quantification cycle (Cq) to define pathogen detection was set to <35. *Shigella spp*. were detected by the *ipaH* gene, as previously outlined.^2^

### Modeled vaccine impacts

We estimated the impact of vaccines on the following outcomes. First, *Shigella* diarrhea was defined as diarrhea episodes with an episode-specific attributable fraction for *Shigella* (AF*e*) >0.5, regardless of other pathogens detected. AF*e*s were calculated as 1 - (1/OR*e*), where OR*e* was the pathogen-specific and quantity-specific odds ratio (OR) from a generalized linear mixed model associating pathogen quantity with diarrhea.^2^ Second, severe *Shigella* diarrhea was defined as *Shigella* diarrhea with a modified Vesikari score >6.^15^ Third, the number of severe diarrhea episodes of any etiology was defined as diarrhea due to any cause with a modified Vesikari score >6. Fourth, diarrhea episodes overall included any etiology and severity.

For vaccine impacts on antibiotic use, we focused on F/Ms as specific drug classes of interest and additionally assessed any antibiotic use. Each diarrhea episode was considered treated with antibiotics if antibiotics were taken during any day of the illness episode. Antibiotic courses overall were defined by antibiotic courses given to the child for any reason, as previously determined.^5^ Antibiotic courses were separated by two antibiotic-free days. Antibiotic exposures to bystander pathogens (i.e., pathogens present at the time of antibiotic treatment, but that did not cause the illness that was treated) were defined by linking each antibiotic course to the most recent stool sample collected in the preceding 30 days. Any bacterial pathogens (atypical enteropathogenic *Escherichia coli* (*E.coli*), *Campylobacter*, enteroaggregative *E. coli*, enterotoxigenic *E. coli*, and typical enteropathogenic *E. coli*) detected in the linked stool were assumed to be bystander pathogens during the antibiotic course.^5^ Antibiotic exposures to bystander pathogens were attributed to the treatment of *Shigella* if the antibiotic course was given during a diarrhea episode with a *Shigella* AFe > 0.5.

### Vaccination scenarios

The characteristics of our simulated *Shigella* vaccine were modeled after those outlined in the WHO’s PPC for a *Shigella* vaccine^1^ and those from vaccines currently in the pipeline. We considered one- and two-dose *Shigella* vaccines with multiple potential vaccine dosing schedules: a one-dose vaccine with administration at 6 months or 9 months, and two dose vaccines with administration at 6 & 9 months, 9 & 12 months, and 12 & 15 months (Table 1). Vaccine efficacy 14 days after the second dose against severe *Shigella* diarrhea was simulated at 60% or 80% in separate scenarios. Efficacy against non-severe episodes was 40% and 60%, respectively. Vaccine efficacy between the first dose up to 14 days after the second dose was half that which was applied 14 days after the second dose. (Table 1).

**Table 1.** *Shigella* vaccination scenarios simulated in the MAL-ED dataset, including dosing schedules, efficacies, and inclusion of indirect and boosting protection.

| Scenario | Dosing schedule | *Shigella*<br>diarrhea<br>Severity | Efficacy 14<br>days after<br>1<sup>st</sup> dose | Efficacy<br>14 days<br>after 2<sup>nd</sup><br>dose | Indirect<br>effect | Boosting effect after 1<sup>st</sup><br>dose, before 2<sup>nd</sup> dose | Boosting effect<br>after 2<sup>nd</sup> dose |
| --- | --- | --- | --- | --- | --- | --- | --- |
| 0 | No vaccine | -- | -- | -- | -- | -- | -- |
| 1 | 1<sup>st</sup>: 6 months | Severe | 60% | -- | 20% | +20% (80% VE) | -- |
|  |  | Non-severe | 40% | -- | 20% | +20% (60% VE) | -- |
| 2 | 1<sup>st</sup>: 9 months | Severe | 60% | -- | 20% | +20% (80% VE) | -- |
|  |  | Non-severe | 40% | -- | 20% | +20% (60% VE) | -- |
| 3 | 1<sup>st</sup>: 6 months<br>2<sup>nd</sup>: 9 months | Severe | 30% | 60% | 20% | +10% (40% VE) | +20% (80% VE) |
|  |  | Non-severe | 20% | 40% | 20% | +10% (30% VE) | +20% (60% VE) |
| 4 | 1<sup>st</sup>: 9 months<br>2<sup>nd</sup>: 12 months | Severe | 30% | 60% | 20% | +10% (40% VE) | +20% (80% VE) |
|  |  | Non-severe | 20% | 40% | 20% | +10% (30% VE) | +20% (60% VE) |
| 5 | 1<sup>st</sup>: 12 months<br>2<sup>nd</sup>: 15 months | Severe | 30% | 60% | 20% | +10% (40% VE) | +20% (80% VE) |
|  |  | Non-severe | 20% | 40% | 20% | +10% (30% VE) | +20% (60% VE) |
| 6 | 1<sup>st</sup>: 6 months | Severe | 80% | -- | 20% | +20% (100% VE) | -- |
|  |  | Non-severe | 60% | -- | 20% | +20% (80% VE) | -- |
| 7 | 1<sup>st</sup>: 9 months | Severe | 80% | -- | 20% | +20% (100% VE) | -- |
|  |  | Non-severe | 60% | -- | 20% | 20% (80% VE) | -- |
| 8 | 1<sup>st</sup>: 6 months<br>2<sup>nd</sup>: 9 months | Severe | 40% | 80% | 20% | +10% (50% VE) | +20% (100% VE) |
|  |  | Non-severe | 30% | 60% | 20% | +10% (40% VE) | +20% (80% VE) |
| 9 | 1<sup>st</sup>: 9 months<br>2<sup>nd</sup>: 12 months | Severe | 40% | 80% | 20% | +10% (50% VE) | +20% (100% VE) |
|  |  | Non-severe | 30% | 60% | 20% | +10% (40% VE) | +20% (80% VE) |
| 10 | 1<sup>st</sup>: 12 months<br>2<sup>nd</sup>: 15 months | Severe | 40% | 80% | 20% | +10% (50% VE) | +20% (100% VE) |
|  |  | Non-severe | 30% | 60% | 20% | +10% (40% VE) | +20% (80% VE) |

For scenarios that assumed the vaccine would produce indirect protection, we randomly selected 20% of *Shigella* diarrhea episodes that occurred in children under the age of the first dose of vaccine administration to be prevented by the vaccine. These simulated levels of indirect protection were based on what was observed with the Vi-tetanus toxoid conjugate vaccine in Bangladesh.^17^ For instance, under a scenario with vaccine doses administered at 9 & 12 months, 20% of diarrhea episodes occurring in children under the age of 9 months were randomly prevented. For scenarios that assumed the vaccine would perform better among children previously exposed to *Shigella* (i.e., boosting protection), efficacy was increased by an absolute 20% for *Shigella* diarrhea episodes that occurred in children who had a *Shigella* infection prior to administration of the first dose of the vaccine. For example, in the 9-& 12-month dosing vaccine scenario with 60% efficacy with 20% boosting effects, if a child was infected with *Shigella* prior to 9 months of age, their allocated efficacy went from 60% to 80%.

Our primary simulated vaccine was a leaky vaccine,^12, 13^ for which we applied a constant proportional reduction (equal to the efficacy) across all diarrhea episodes and antibiotic courses (i.e., the vaccine prevented 60% of all diarrhea episodes or antibiotic courses). In a sensitivity analysis, we simulated an all-or-nothing vaccine,^12, 13^ for which we selected a subset of children who were vaccine responders at random, the size of which was defined by vaccine efficacy, and prevented all diarrhea episodes and antibiotic courses in those children (i.e., all episodes among 60% of children were prevented).

Results reported primarily in the text correspond to a leaky vaccine with two doses at 9 and 12 months with 60% efficacy since these characteristics may be the most realistic among the range of acceptable parameters outlined in the WHO’s PPC.^1^ Results from all other vaccination scenarios are described in the tables and figures.

### Statistical analysis

To estimate the incidence of each diarrhea and antibiotic outcome defined above expected under each vaccination scenario, we performed Monte Carlo simulations using random sampling with replacement of children to a sample size of 50,000. For each simulation, we randomly selected *Shigella* diarrhea episodes from these children to be prevented by a probability equal to vaccine efficacy and calculated the incidence of each outcome excluding prevented episodes. In a no-vaccine scenario, no episodes were selected to be prevented. Estimates and confidence intervals were estimated by the median, 2.5^th^ and 97.5^th^ percentiles of 1,000 iterations of this procedure. To quantify the expected reductions in the outcomes listed above, we estimated absolute and relative differences and the percent change between each vaccine scenario and the no vaccine scenario. All statistical analyses were performed via R software, version 4.0.2 (Foundation for Statistical Computing).

### Ethics approvals and data availability

This study involves human participants. For the parent study, ethical approval was obtained from the Institutional Review Boards at the University of Virginia School of Medicine (Charlottesville, USA) (14595) and at each of the participating research sites: Ethical Review Committee, ICDDR,B (Bangladesh); Committee for Ethics in Research, Universidade Federal do Ceara; National Ethical Research Committee, Health Ministry, Council of National Health (Brazil); Institutional Review Board, Christian Medical College, Vellore; Health Ministry Screening Committee, Indian Council of Medical Research (India); Institutional Review Board, Institute of Medicine, Tribhuvan University; Ethical Review Board, Nepal Health Research Council; Institutional Review Board, Walter Reed Army Institute of Research (Nepal); Institutional Review Board, Johns Hopkins University; PRISMA Ethics Committee; Health Ministry, Loreto (Peru); Ethical Review Committee, Aga Khan University (Pakistan); Health, Safety and Research Ethics Committee, University of Venda; Department of Health and Social Development, Limpopo Provincial Government (South Africa); Medical Research Coordinating Committee, National Institute for Medical Research; Chief Medical Officer, Ministry of Health and Social Welfare (Tanzania). For the current study, we obtained ethical approval at the University of Virginia School of Medicine (Charlottesville, USA) (22398) and Emory University (Atlanta, USA) (STUDY00003285). Participants gave informed consent to participate in the study before taking part. The statistical analysis plan is available at osf.io/3asxh. Deidentified participant data from the MAL-ED study is publicly available at ClinEpiDB.org.

### Role of the funding source

The funders of this study did not have any role in the study design, collection, analysis, interpretation of the data, writing of the report, nor in the decision to submit the paper for publication.

## RESULTS

These analyses included 1715 children, of which 83% (n=1427) had at least one *Shigella* infection during their first two years of life (Table 2). There were 273.8 diarrhea episodes of any severity per 100 child-years (n=9392) and 30.6 *Shigella* diarrhea episodes per 100 child years (n=754). Caregivers reported 457.6 courses per 100 child years of antibiotics (n=15697), amongst which 110.1 courses per 100 child years (n=3775) were to treat diarrhea episodes of any etiology and of which 16.3 courses per 100 child years were attributable to *Shigella* diarrhea (n=427). Bystander pathogens had 646.1 (n=22161) and 32.9 (n=750) exposures to antibiotics per 100 child years resulting from any antibiotic use and resulting from the treatment of *Shigella*, respectively.

**Table 2.**
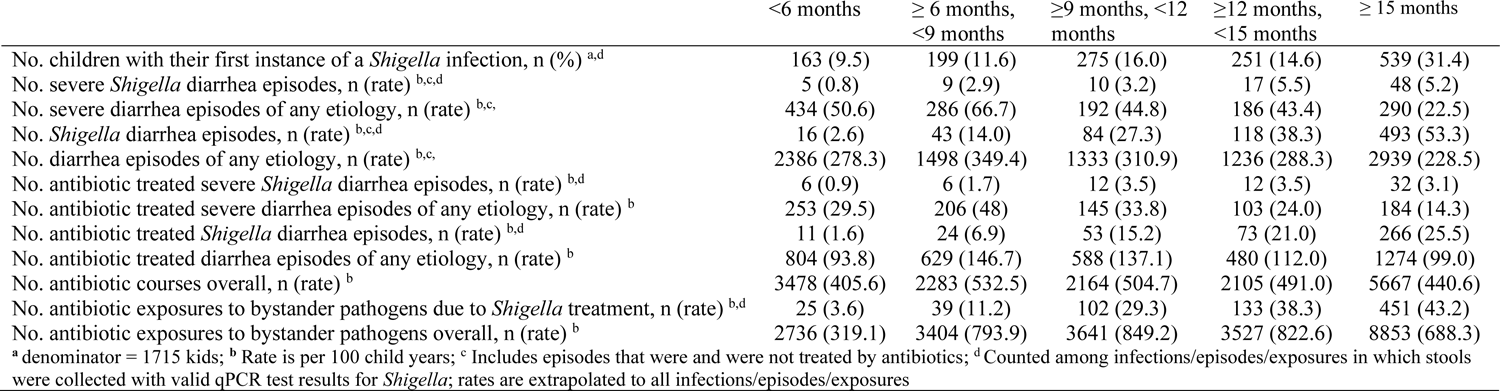
Diarrhea episodes, antibiotic use, and bystander pathogen exposures to antibiotics among 1715 children enrolled in the MAL-ED cohort.

### Prevention of diarrhea

A leaky *Shigella* vaccine given at 9 & 12 months with 60% efficacy would be expected to prevent 1.7 (95% CI: 1.3, 2.1) severe *Shigella* diarrhea episodes and 11.0 (95% CI: 10.0, 11.9) *Shigella* diarrhea episodes of any severity per 100 child years (Table 3), which corresponds to a 46.5% reduction in severe shigellosis episodes and a 35.9% reduction in shigellosis episodes of any severity (Figure 1, Supp Table 1). While the vaccine would reduce the same number of severe and all diarrhea episodes due to any etiology, the percent reductions would be smaller, at 3.7% for severe diarrhea episodes of any etiology and 4.0% for diarrhea episodes of any etiology (Table 3, Figure 1, Supp Table 1).

**Figure 1.**
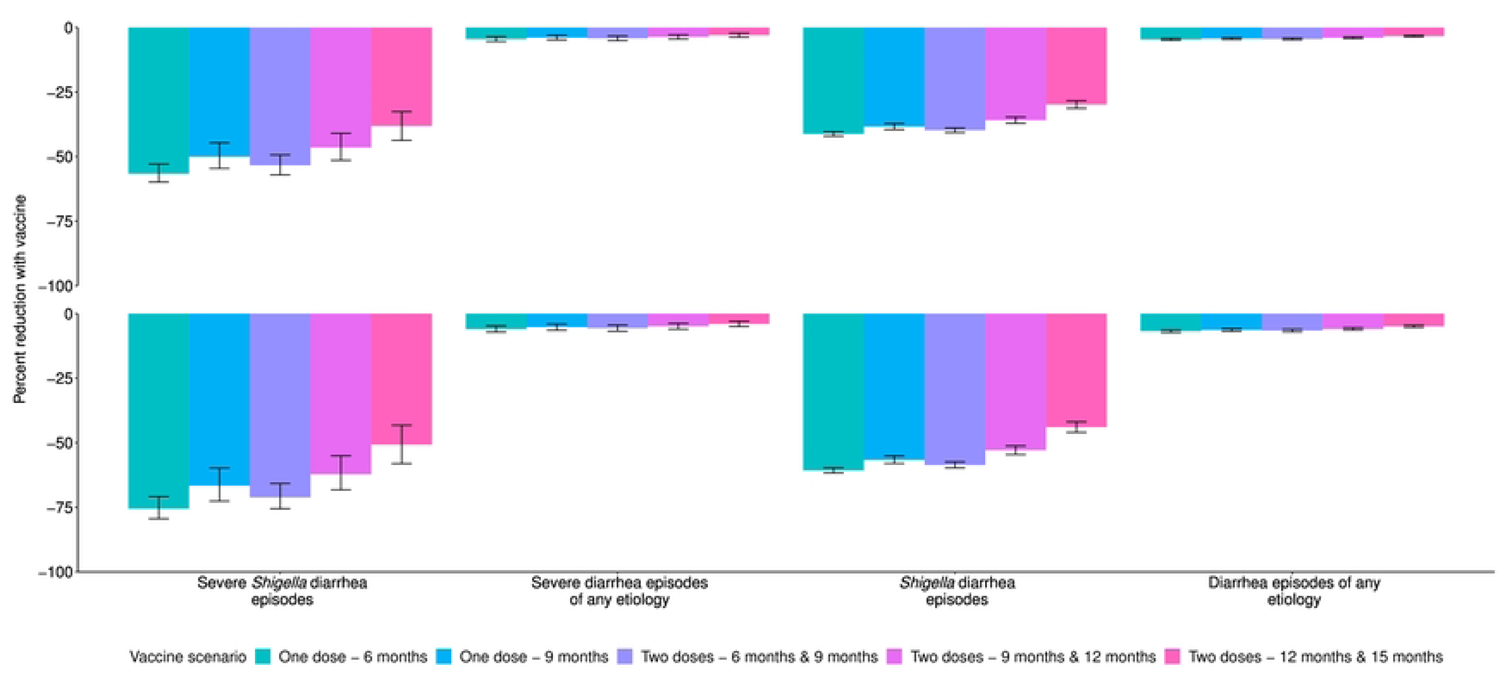
Percent reductions in diarrhea outcomes among five vaccine scenarios with 60% (A) and 80% (B) full vaccine efficacies and no indirect or boosting protection.

**Table 3.** Absolute and relative differences in diarrhea outcomes among five vaccine scenarios with 60% and 80% full vaccine efficacies and no indirect or boosting protection.

| Vaccine scenario and efficacy outcome | Absolute difference<br>(cases per 100 child-years) |  | Relative difference |  |
| --- | --- | --- | --- | --- |
|  | 60% VE<br>(95% CI) | 80% VE<br>(95% CI) | 60% VE<br>(95% CI) | 80% VE<br>(95% CI) |
| One dose - 6 months |  |  |  |  |
| Severe *Shigella* diarrhea episodes | 2.0 (1.6, 2.5) | 2.7 (2.2, 3.4) | 0.43 (0.40, 0.47) | 0.24 (0.21, 0.29) |
| Severe diarrhea episodes of any etiology | 2.0 (1.6, 2.5) | 2.7 (2.2, 3.4) | 0.96 (0.95, 0.96) | 0.94 (0.93, 0.95) |
| *Shigella* diarrhea episodes | 12.6 (11.6, 13.7) | 18.6 (17.1, 20.2) | 0.59 (0.58, 0.60) | 0.39 (0.38, 0.40) |
| Diarrhea episodes of any etiology | 12.6 (11.6, 13.7) | 18.6 (17.1, 20.2) | 0.95 (0.95, 0.96) | 0.93 (0.93, 0.94) |
| One dose - 9 months |  |  |  |  |
| Severe *Shigella* diarrhea episodes | 1.8 (1.4, 2.3) | 2.4 (1.8, 3.0) | 0.50 (0.45, 0.55) | 0.33 (0.27, 0.40) |
| Severe diarrhea episodes of any etiology | 1.8 (1.4, 2.3) | 2.4 (1.8, 3.0) | 0.96 (0.95, 0.97) | 0.95 (0.94, 0.96) |
| *Shigella* diarrhea episodes | 11.8 (10.8, 12.7) | 17.3 (15.9, 18.8) | 0.62 (0.60, 0.63) | 0.43 (0.42, 0.45) |
| Diarrhea episodes of any etiology | 11.8 (10.8, 12.7) | 17.3 (15.9, 18.8) | 0.96 (0.95, 0.96) | 0.94 (0.93, 0.94) |
| Two doses - 6 months & 9 months |  |  |  |  |
| Severe *Shigella* diarrhea episodes | 1.9 (1.5, 2.4) | 2.6 (2.0, 3.2) | 0.47 (0.43, 0.51) | 0.29 (0.24, 0.34) |
| Severe diarrhea episodes of any etiology | 1.9 (1.5, 2.4) | 2.6 (2.0, 3.2) | 0.96 (0.95, 0.97) | 0.94 (0.93, 0.96) |
| *Shigella* diarrhea episodes | 12.2 (11.2, 13.2) | 17.9 (16.5, 19.4) | 0.60 (0.59, 0.61) | 0.41 (0.40, 0.43) |
| Diarrhea episodes of any etiology | 12.2 (11.2, 13.2) | 17.9 (16.5, 19.4) | 0.96 (0.95, 0.96) | 0.93 (0.93, 0.94) |
| Two doses - 9 months & 12 months |  |  |  |  |
| Severe *Shigella* diarrhea episodes | 1.7 (1.3, 2.1) | 2.2 (1.7, 2.8) | 0.53 (0.49, 0.59) | 0.38 (0.32, 0.45) |
| Severe diarrhea episodes of any etiology | 1.7 (1.3, 2.1) | 2.2 (1.7, 2.8) | 0.96 (0.95, 0.97) | 0.95 (0.94, 0.96) |
| *Shigella* diarrhea episodes | 11.0 (10.0, 11.9) | 16.2 (14.8, 17.6) | 0.64 (0.63, 0.65) | 0.47 (0.45, 0.49) |
| Diarrhea episodes of any etiology | 11.0 (10.0, 11.9) | 16.2 (14.8, 17.6) | 0.96 (0.96, 0.96) | 0.94 (0.94, 0.95) |
| Two doses - 12 months & 15 months |  |  |  |  |
| Severe *Shigella* diarrhea episodes | 1.4 (1.0, 1.7) | 1.8 (1.4, 2.3) | 0.62 (0.56, 0.67) | 0.49 (0.42, 0.57) |
| Severe diarrhea episodes of any etiology | 1.4 (1.0, 1.7) | 1.8 (1.4, 2.3) | 0.97 (0.96, 0.98) | 0.96 (0.95, 0.97) |
| *Shigella* diarrhea episodes | 9.1 (8.3, 10.0) | 13.4 (12.2, 14.7) | 0.70 (0.69, 0.72) | 0.56 (0.54, 0.58) |
| Diarrhea episodes of any etiology | 9.1 (8.3, 10.0) | 13.4 (12.2, 14.7) | 0.97 (0.96, 0.97) | 0.95 (0.95, 0.96) |

The 11.0 (95% CI: 10.0, 11.9) prevented *Shigella* diarrhea episodes per 100 child years increased slightly to 11.5 (95% CI: 10.6, 12.5) when analyses further allowed for 20% indirect protection (Supp Table 2). This same vaccine with 20% boosting protection and no indirect protection would prevent 12.8 (95% CI: 11.7, 14.0) *Shigella* diarrhea episodes (Supp Table 3). Together, a vaccine with direct effects plus indirect and boosting protection effects would prevent 13.4 (95% CI: 12.3, 14.5) *Shigella* diarrhea episodes per 100 child years (Supp Table 4), which equates to a 43.7% reduction in *Shigella* diarrhea episodes and a 54.5% reduction in severe *Shigella* diarrhea episodes (Figure 2, Supp Table 5).

**Figure 2.**
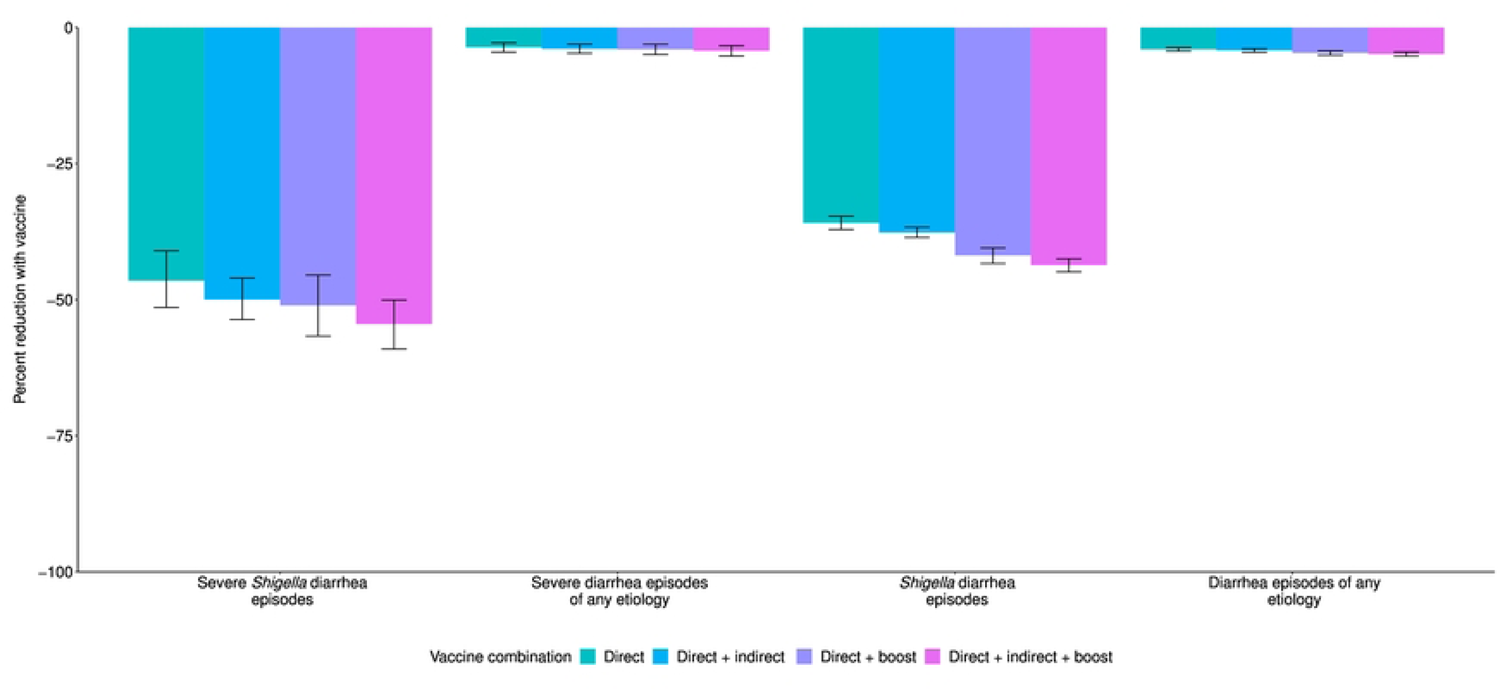
Percent reductions in diarrhea outcomes with the addition of indirect and boosting protection among the 9- and 12-month vaccine dosing scenario with 60% full vaccine efficacy.

**Table 4.** Absolute and relative differences in fluoroquinolone/macrolide (F/M) outcomes in among five vaccine scenarios with 60% and 80% full vaccine efficacies and no indirect or boosting protection.

| Vaccine scenario and efficacy outcome | Absolute difference<br>(cases per 100 child-years) |  | Relative difference |  |
| --- | --- | --- | --- | --- |
|  | 60% VE (95% CI) | 80% VE (95% CI) | 60% VE (95% CI) | 80% VE (95% CI) |
| One dose - 6 months |  |  |  |  |
| F/M treated severe *Shigella* diarrhea episodes | 0.5 (0.3, 0.8) | 0.7 (0.4, 1.0) | 0.45 (0.38, 0.56) | 0.27 (0.19, 0.41) |
| F/M treated severe diarrhea episodes of any etiology | 0.5 (0.3, 0.8) | 0.7 (0.4, 1.0) | 0.92 (0.89, 0.95) | 0.90 (0.85, 0.94) |
| F/M treated *Shigella* diarrhea episodes | 3.4 (2.9, 4.0) | 5.1 (4.3, 5.9) | 0.58 (0.57, 0.60) | 0.39 (0.37, 0.41) |
| F/M treated diarrhea episodes of any etiology | 3.4 (2.9, 4.0) | 5.1 (4.3, 5.9) | 0.91 (0.90, 0.92) | 0.87 (0.85, 0.88) |
| F/M courses overall | 3.4 (2.9, 4.0) | 5.1 (4.3, 5.9) | 0.97 (0.96, 0.97) | 0.95 (0.95, 0.96) |
| F/M exposures to bystander pathogens due to *Shigella* treatment | 7.2 (5.9, 8.4) | 10.6 (8.8, 12.5) | 0.59 (0.57, 0.61) | 0.39 (0.37, 0.42) |
| F/M exposures to bystander pathogens overall | 7.2 (5.9, 8.4) | 10.6 (8.8, 12.5) | 0.96 (0.96, 0.97) | 0.95 (0.94, 0.95) |
| One dose - 9 months |  |  |  |  |
| F/M treated severe *Shigella* diarrhea episodes | 0.5 (0.3, 0.7) | 0.6 (0.4, 1.0) | 0.49 (0.41, 0.62) | 0.33 (0.22, 0.49) |
| F/M treated severe diarrhea episodes of any etiology | 0.5 (0.3, 0.7) | 0.6 (0.4, 1.0) | 0.93 (0.90, 0.96) | 0.91 (0.86, 0.95) |
| F/M treated *Shigella* diarrhea episodes | 3.2 (2.7, 3.8) | 4.8 (4, 5.6) | 0.61 (0.59, 0.63) | 0.42 (0.40, 0.45) |
| F/M treated diarrhea episodes of any etiology | 3.2 (2.7, 3.8) | 4.8 (4, 5.6) | 0.91 (0.90, 0.92) | 0.87 (0.86, 0.89) |
| F/M courses overall | 3.2 (2.7, 3.8) | 4.8 (4, 5.6) | 0.97 (0.97, 0.97) | 0.96 (0.95, 0.96) |
| F/M exposures to bystander pathogens due to *Shigella* treatment | 6.7 (5.6, 8.0) | 9.9 (8.3, 11.7) | 0.61 (0.59, 0.64) | 0.43 (0.40, 0.46) |
| F/M exposures to bystander pathogens overall | 6.7 (5.6, 8.0) | 9.9 (8.3, 11.7) | 0.97 (0.96, 0.97) | 0.95 (0.94, 0.96) |
| Two doses - 6 months & 9 months |  |  |  |  |
| F/M treated severe *Shigella* diarrhea episodes | 0.5 (0.3, 0.8) | 0.7 (0.4, 1.0) | 0.48 (0.4, 0.58) | 0.30 (0.21, 0.45) |
| F/M treated severe diarrhea episodes of any etiology | 0.5 (0.3, 0.8) | 0.7 (0.4, 1.0) | 0.93 (0.89, 0.96) | 0.90 (0.86, 0.94) |
| F/M treated *Shigella* diarrhea episodes | 3.4 (2.8, 3.9) | 4.9 (4.2, 5.8) | 0.60 (0.58, 0.61) | 0.40 (0.38, 0.43) |
| F/M treated diarrhea episodes of any etiology | 3.4 (2.8, 3.9) | 4.9 (4.2, 5.8) | 0.91 (0.90, 0.92) | 0.87 (0.85, 0.88) |
| F/M courses overall | 3.4 (2.8, 3.9) | 4.9 (4.2, 5.8) | 0.97 (0.96, 0.97) | 0.95 (0.95, 0.96) |
| F/M exposures to bystander pathogens due to *Shigella* treatment | 6.9 (5.8, 8.2) | 10.2 (8.6, 12.0) | 0.60 (0.58, 0.62) | 0.41 (0.39, 0.44) |
| F/M exposures to bystander pathogens overall | 6.9 (5.8, 8.2) | 10.2 (8.6, 12.0) | 0.97 (0.96, 0.97) | 0.95 (0.94, 0.96) |
| Two doses - 9 months & 12 months |  |  |  |  |
| F/M treated severe *Shigella* diarrhea episodes | 0.5 (0.2, 0.7) | 0.6 (0.3, 0.9) | 0.52 (0.42, 0.63) | 0.35 (0.24, 0.50) |
| F/M treated severe diarrhea episodes of any etiology | 0.5 (0.2, 0.7) | 0.6 (0.3, 0.9) | 0.93 (0.90, 0.96) | 0.91 (0.86, 0.95) |
| F/M treated *Shigella* diarrhea episodes | 3.1 (2.6, 3.7) | 4.5 (3.8, 5.4) | 0.63 (0.61, 0.65) | 0.45 (0.42, 0.48) |
| F/M treated diarrhea episodes of any etiology | 3.1 (2.6, 3.7) | 4.5 (3.8, 5.4) | 0.92 (0.91, 0.93) | 0.88 (0.86, 0.89) |
| F/M courses overall | 3.1 (2.6, 3.7) | 4.5 (3.8, 5.4) | 0.97 (0.97, 0.98) | 0.96 (0.95, 0.96) |
| F/M exposures to bystander pathogens due to *Shigella* treatment | 6.3 (5.2, 7.5) | 9.3 (7.7, 11.1) | 0.64 (0.61, 0.66) | 0.47 (0.43, 0.50) |
| F/M exposures to bystander pathogens overall | 6.3 (5.2, 7.5) | 9.3 (7.7, 11.1) | 0.97 (0.96, 0.97) | 0.95 (0.95, 0.96) |
| Two doses - 12 months & 15 months |  |  |  |  |
| F/M treated severe *Shigella* diarrhea episodes | 0.4 (0.2, 0.7) | 0.5 (0.3, 0.9) | 0.57 (0.47, 0.69) | 0.42 (0.29, 0.58) |
| F/M treated severe diarrhea episodes of any etiology | 0.4 (0.2, 0.7) | 0.5 (0.3, 0.9) | 0.94 (0.91, 0.97) | 0.92 (0.87, 0.96) |
| F/M treated *Shigella* diarrhea episodes | 2.6 (2.2, 3.1) | 3.9 (3.2, 4.6) | 0.68 (0.66, 0.71) | 0.53 (0.50, 0.57) |
| F/M treated diarrhea episodes of any etiology | 2.6 (2.2, 3.1) | 3.9 (3.2, 4.6) | 0.93 (0.92, 0.94) | 0.90 (0.88, 0.91) |
| F/M courses overall | 2.6 (2.2, 3.1) | 3.9 (3.2, 4.6) | 0.98 (0.97, 0.98) | 0.96 (0.96, 0.97) |
| F/M exposures to bystander pathogens due to *Shigella* treatment | 5.3 (4.3, 6.4) | 7.8 (6.4, 9.4) | 0.70 (0.67, 0.73) | 0.55 (0.51, 0.59) |
| F/M exposures to bystander pathogens overall | 5.3 (4.3, 6.4) | 7.8 (6.4, 9.4) | 0.97 (0.97, 0.98) | 0.96 (0.95, 0.97) |

### Prevention of antibiotic use

A two-dose *Shigella* vaccine given at 9 and 12 months with 60% vaccine efficacy could prevent 0.5 (95% CI: 0.2, 0.7) F/M treated severe *Shigella* diarrhea episodes (48.4% reduction), 3.1 (95% CI: 2.6, 3.7) F/M treated *Shigella* episodes (37.3% reduction), and 6.3 (95% CI: 5.2, 7.5) F/M exposures to bystander pathogens due to *Shigella* treatment (36.2% reduction) per 100 child years (Table 4, Figure 3, Supp Table 6). However, this vaccine would reduce overall F/M uses and overall F/M exposures to bystander pathogens by only 2.9% and 3.2%, respectively (Supp Table 6). The 3.1 (95% CI: 2.6, 3.7) prevented F/M treated *Shigella* diarrhea episodes per 100 child years increased to 3.2 (95% CI: 2.7, 3.7) prevented episodes with added indirect protection effects (Supp Table 7), to 3.6 (95% CI: 3.0, 4.3) with added boosting effects (Supp Table 8), and to 3.7 (95% CI: 3.1, 4.4) when both indirect effects and boosting effects were added to the direct effects (Supp Table 9). When indirect and boosting effects were added to the direct effects, there were slight increases in percent reductions of all metrics: F/M treated *Shigella* diarrhea episodes (37.3% to 45.1%), F/M courses overall (2.9% to 3.5%), F/M exposures to bystander pathogens due to *Shigella* treatment (36.2% to 44.6%), and F/M exposures to bystander pathogens overall (3.2% to 3.9%) (Figure 4, Supp Table 10).

**Figure 3.**
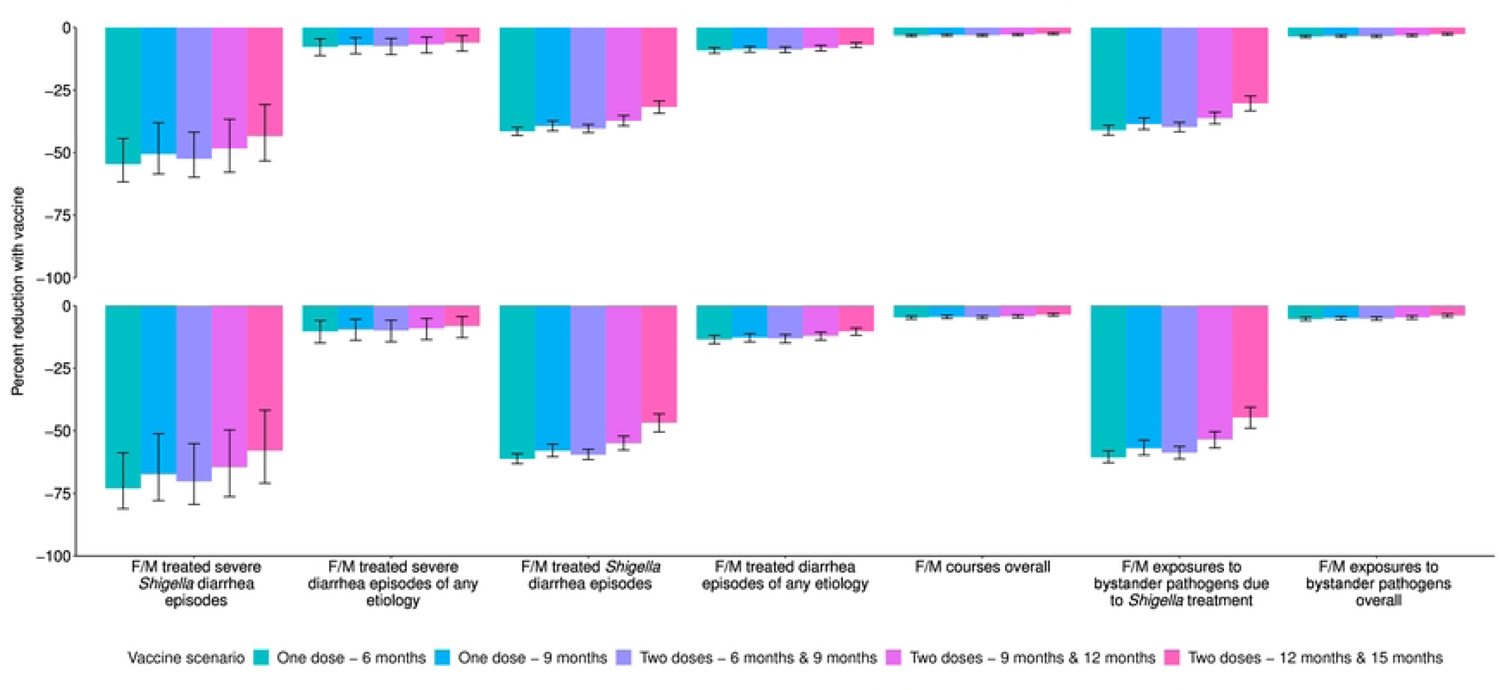
Percent reductions in fluroquinolone and macrolide (F/M) use outcomes among five vaccine scenarios with 60% (A) and 80% (B) full vaccine efficacies and no indirect or boosting protection.

**Figure 4.**
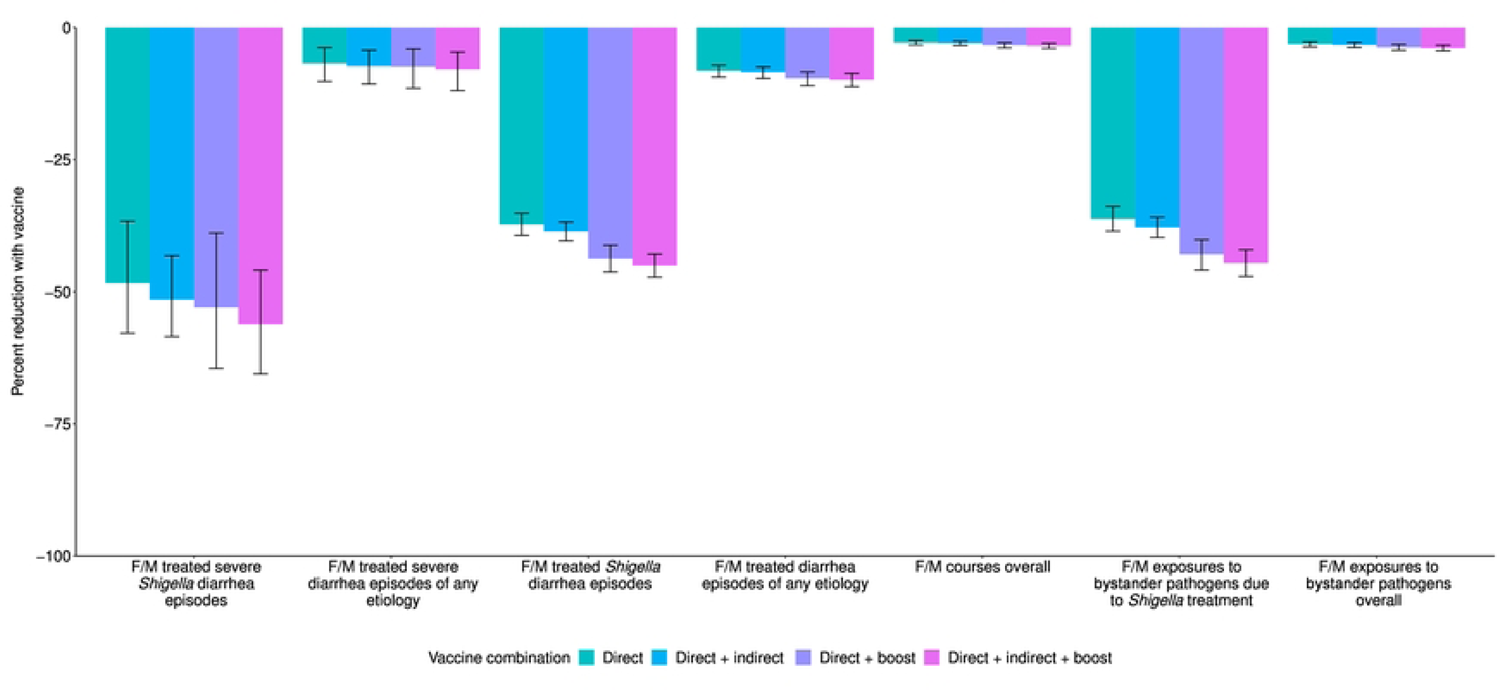
Percent reductions in fluroquinolone and macrolide (F/M) use outcomes with the addition of indirect and boosting protection among the 9- and 12-month vaccine dosing scenario with 60% full vaccine efficacy.

While a two-dose *Shigella* vaccine given at 9 and 12 months with 60% vaccine efficacy would prevent more instances of antibiotic use overall than of F/M specifically, the percent reductions in overall antibiotic use were smaller than what was observed with F/M use (Supp Table 11, Supp Figure 1, Supp Table 12). In this scenario, 1.1 (95% CI: 0.7, 1.4) courses of antibiotic treated severe *Shigella* diarrhea episodes (41.3% reduction), 5.8 (95% CI: 5.2, 6.6) antibiotic treated *Shigella* diarrhea episodes (35.6% reduction), and 11.2 (95% CI: 9.7, 12.9) antibiotic exposures to bystander pathogens due to *Shigella* treatment (35.0% reduction) per 100 child years were prevented (Supp Table 11, Supp Figure 2, Supp Table 12). However, there was only a 1.0% and 1.2% reduction in overall antibiotic uses and overall exposures to bystander pathogens, respectively (Supp Table 12). Similar to what was observed with F/M, the addition of indirect and boosting effects onto the direct effects minimally increased the number of prevented outcomes (Supp Table 13-16, Supp Figure 2).

### All-or-nothing vaccine compared to leaky vaccine

A *Shigella* vaccine given at 9 & 12 months that fully protected 60% of children (i.e., an all-or-nothing vaccine) prevented the same number of severe *Shigella* diarrhea episodes (1.7 (95% CI: 1.3, 2.1) episodes per 100 child years) as the leaky vaccine (i.e., prevented 60% of episodes across children) (Table 3, Supp Table 17). However, the all-or-nothing vaccine prevented more *Shigella* diarrhea episodes of any severity compared to the leaky vaccine (15.6 (95% CI: 14.3, 17.0) vs. 11.0 (95% CI: 10.0, 11.9) episodes per 100 child years) (Table 3, Supp Table 17). Similar results were found for the antibiotic outcomes, such that an all-or-nothing vaccine would be expected to prevent more of all outcomes except the severe outcomes, for which it would prevent the same number as a leaky vaccine (Supp Table 18, Supp Table 19).

The corresponding results for the other vaccine scenarios listed in Table 1 are displayed in Figures 1-2, Tables 3-4, Supp Figure 2, Supp Tables 1-4, 6-9, 11-15, 17-19). In general, the earlier the vaccine is given, the greater the expected reduction in diarrhea episodes and antibiotic use. Additionally, the single dose vaccines were more efficacious than the two-dose vaccines initiated at the same time (e.g., one-dose at 9 months vs. two-doses at 9 and 12 months) since the full efficacy was achieved at an earlier age with the single dose vaccines.

## DISCUSSION

A leaky *Shigella* vaccine administered at 9 and 12 months with 60% vaccine efficacy could provide a substantial reduction in severe *Shigella* diarrhea episodes, *Shigella* diarrhea episodes of any severity, and F/M treated *Shigella* diarrhea episodes. However, given the multitude of causes of diarrhea and antibiotic use in this population, the expected reductions in all-cause diarrhea and antibiotic use overall were modest (<5%). While single-dose vaccines and vaccines given at younger ages would prevent more diarrhea and antibiotic use, none of the vaccine candidates in clinical development meet those criteria.

Incorporation of indirect protection and boosting only slightly increased the number of diarrhea episodes and antibiotic courses expected to be preventable, suggesting these nuances will not be major determinants of vaccine success. A *Shigella* vaccine will likely fall somewhere on the continuum between a leaky (i.e., prevention of a fraction of episodes in all children) and all-or-nothing (i.e., prevention of all episodes in a subset of children who are vaccine responders) vaccine.^12, 13^ As expected, the all-or-nothing vaccine effects were larger than the leaky vaccine effects for outcomes that were not limited to severe diarrhea because an all-or-nothing vaccine would provide complete protection in a subset of individuals regardless of disease severity. However, our analysis was limited by assuming that vaccine responders were a random subset of the population. It may be more likely that vaccine responders would be expected to be at lower risk of shigellosis even in the absence of vaccine. Furthermore, while our results estimate the upper limit of the potential benefit of a *Shigella* vaccine since we assumed 100% vaccine coverage, it is likely that vaccine coverage would be lower in a real-world setting.

The absolute reductions in F/M use achieved by a *Shigella* vaccine accounted for roughly half the achievable reduction of all antibiotic use. However, there were greater percent reductions in F/M use compared to all antibiotic use since F/Ms are often targeted for diarrhea treatment, and specifically for dysentery presumed to be shigellosis. F/M use has been associated with resistance in these drug classes,^5, 8, 18^ suggesting reductions in use achievable by a *Shigella* vaccine could limit drug-resistant shigellosis as well as the development of resistance in other enteric bacteria through reductions in bystander exposure. Our predicted reductions in all antibiotic and F/M use could be underestimates if suspicion of *Shigella* is the main reason for treating diarrhea regardless of etiology, such that treatment rates also decline for other diarrhea etiologies after *Shigella* incidence is known to have been substantially reduced by the vaccine. As *Shigella* vaccines are evaluated in large Phase III trials, data on antibiotic treatment should be carefully collected such that the impact of the vaccine on antibiotic use can be measured.^1, 19^ To quantify this impact, it will be important for vaccine effectiveness to be estimated against less-severe disease endpoints which account for the bulk of antibiotic use.

The vaccine scenarios modeled are in line with the WHO PPC guidance. However, some of the current vaccines in the pipeline require more doses. There is one phase III *Shigella* vaccine (ZF0901 (Beijing Zhifei Lvzhu Biopharmaceutical Co., Ltd.))^20^ and three phase IIA *Shigella* vaccines (Shigella4V (Limmatech AG)^21^, altSonflex1-2-3 (GVGH),^22^ and GlycoShig3 (Institut Pasteur)^23^) in the pipeline.^3^ ZF0901 is 3 doses for infants ages 3-6 months and 2 doses for those aged 6-12 months. Shigella4V is 3 doses for infants (8 months +/-1 month) and children (2-5 years). altSonflex1-2-3 is 3 doses for infants 9 months of age and changes to 2 doses for children 24-59 months. GlycoShig3 is 3 doses infants (9 months +/-1 month) and children (2-5 years).

For our age group of interest (children under two years of age), the aforementioned vaccines would all require 2-3 doses. Our estimates of the expected reductions in outcomes would apply to a three-dose vaccine where the full efficacy is achieved after two doses. If full efficacy is not achieved until a third dose, our expected reductions may be overestimated.

A *Shigella* vaccine will likely not be protective against all serotypes, but given the lack of serotyping data, we were unable to simulate the prevention of episodes at the serotype level. However, there is no need for a *Shigella* vaccine to contain all 49 serotypes for broad coverage. *Shigella flexneri* (*S. flexneri*) is the leading cause of endemic diarrhea in LMICs while *Shigella sonnei* (*S. sonnei*) is the leading cause in high income countries.^6^ While there are 15 serotypes for *S. flexneri*, five (2a, 1b, 2b, 3a, 6) accounted for 89% of *S. flexneri* isolates from the Global Enteric Multicenter Study (GEMS).^24^ On a yearly basis, there are minimal changes to the dominant *Shigella* serotypes. Therefore, a quadrivalent vaccine with *S. sonnei* and *S. flexneri 2a, 3a,* and *6* could provide 64% protection against *Shigella* with coverage up to 88% via cross protection.^24^ Our estimates assume 100% cross protection for subtypes not included in the vaccine and therefore may be slightly overestimated depending on the true levels of cross protection observed.

Finally, we did not consider the potential for waning immunity since we only observed outcomes to two years of age. The effects of waning would likely occur more than 6 months after the last vaccine dose, which was outside of our follow-up period for most vaccine scenarios. However, if efficacy wanes substantially before two years of age, our expected reductions may also be overestimated. Since waning may vary by endpoint (i.e., more pronounced waning for less severe disease), this will be another important feature to monitor in trials.

It is important to note that while we targeted efficacies at 60% and 80%, those proportions of outcomes were not prevented at the population level for two reasons: 1) targeted efficacies of 60% and 80% were for severe diarrhea only (as per the PPC) whereas efficacy was lower for non-severe diarrhea and 2) episodes that occur prior to vaccination would not be expected to be prevented. Because severe disease is more common in younger children, the expected relative reductions in severe outcomes were particularly less for vaccine strategies with older ages of administration. However, given the high burden of shigellosis and antibiotic treatment of shigellosis, a *Shigella* vaccine could still make a substantial impact on *Shigella* diarrhea burden in term of absolute reduction in episodes, and have ancillary benefits in the reduction of antibiotic use.

Our estimates provide more realistic expectations for the reductions in diarrhea outcomes at the population-level that could be achieved by *Shigella* vaccines under real world introduction scenarios. A previous modeling study estimated similar absolute reductions in *Shigella* diarrhea episodes under a more limited set of vaccine assumptions that did not account for partial protection after a first dose, herd immunity, or effects on antibiotic use.^25^ Uniquely, we demonstrate that *Shigella* vaccines could provide important reductions in antibiotic use for severe and non-severe *Shigella* diarrheal episodes, and exposures to bystander pathogens due to *Shigella* treatment. Given the high burden of enteric infections and antibiotic use among children in LMICs, the value proposition of a *Shigella* vaccine in this population is strong and substantially augmented by the projected impacts on antibiotic use.

## Data Availability

The statistical analysis plan is available at osf.io/3asxh. Deidentified participant data from the MAL-ED study is publicly available at ClinEpiDB.org.

## ACKNOWLEDGEMENT

This work was supported by Wellcome (219741/Z/19/Z to ETRM). The Etiology, Risk Factors and Interactions of Enteric Infections and Malnutrition and the Consequences for Child Health and Development Project (MAL-ED) was a collaborative project supported by the Bill & Melinda Gates Foundation (OPP1131125), the Foundation for the NIH, the National Institutes of Health, and the Fogarty International Center.

## DATA SHARING

De-identified participant data from the MAL-ED study is publicly available at ClinEpiDB.org after approval of a proposal by the study PIs.

## AUTHORS’ CONTIBUTIONS

SAB and ETRM led data analysis, visualization, interpretation, and writing of the report. JAP-M contributed to data analysis, methodology, and visualization. JAP-M and JAL contributed to interpretation. JL led the development of the laboratory assays. ERH led funding acquisition and administration of the parent study. JAP-M, JAL, JL, and ERH contributed to reviewing/editing the report. ETRM led conceptualization, methodology, and funding acquisition.

## DECLARATION OF INTEREST

We declare no competing interests.

## Notes

### Competing Interest Statement

The authors have declared no competing interest.

### Funding Statement

This work was supported by Wellcome (219741/Z/19/Z to ETRM wellcome.org). The Etiology, Risk Factors and Interactions of Enteric Infections and Malnutrition and the Consequences for Child Health and Development Project (MAL-ED) was a collaborative project supported by the Bill & Melinda Gates Foundation (OPP1131125 gatesfoundation.org), the Foundation for the NIH, the National Institutes of Health, and the Fogarty International Center. The funders had no role in study design, data collection and analysis, decision to publish, or preparation of the manuscript.

### Author Declarations

This study involves human participants. For the parent study, ethical approval was obtained from the Institutional Review Boards at the University of Virginia School of Medicine (Charlottesville, USA) (14595) and at each of the participating research sites: Ethical Review Committee, ICDDR,B (Bangladesh) Committee for Ethics in Research, Universidade Federal do Ceara National Ethical Research Committee, Health Ministry, Council of National Health (Brazil) Institutional Review Board, Christian Medical College, Vellore Health Ministry Screening Committee, Indian Council of Medical Research (India) Institutional Review Board, Institute of Medicine, Tribhuvan University Ethical Review Board, Nepal Health Research Council Institutional Review Board, Walter Reed Army Institute of Research (Nepal) Institutional Review Board, Johns Hopkins University PRISMA Ethics Committee Health Ministry, Loreto (Peru) Ethical Review Committee, Aga Khan University (Pakistan) Health, Safety and Research Ethics Committee, University of Venda Department of Health and Social Development, Limpopo Provincial Government (South Africa) Medical Research Coordinating Committee, National Institute for Medical Research Chief Medical Officer, Ministry of Health and Social Welfare (Tanzania). For the current study, we obtained ethical approval at the University of Virginia School of Medicine (Charlottesville, USA) (22398) and Emory University (Atlanta, USA) (STUDY00003285).

